# Child marriage and adverse reproductive outcomes among young Afghan women: implication for policy and practice in Afghanistan

**DOI:** 10.1101/2021.11.20.21266626

**Authors:** Omid Dadras, Takeo Nakayama

## Abstract

**Background:** Child marriage is defined as marriage before the age of 18 years and it has been linked to several adverse health and social outcomes. This study aimed to explore the prevalence and determinants of child marriage and its association with adverse reproductive outcomes among a nationally representative sample of young Afghan women.

**Methods:** This was a secondary analysis of the 2015 Afghanistan Demographic and Health Survey (DHS) data. Based on the UN recommendation on child marriage study, only women aged 20-24 years old were included. Descriptive statistics and binary logistic regression were employed to determine the distribution of respondents’ characteristics and prevalence and determinants of child marriage. Multivariate logistic models examined the association between child marriage and adverse reproductive outcomes accounting for the sociodemographic factors.

**Results:** An estimated 52% of the Afghan women aged 20-24 married at ages less than 18 years. Poor illiterate women were more likely to marry at early ages (<18). There was a significant negative relationship between child marriage and history of rapid repeat childbirth, delivery by skilled personnel, and institutional delivery. In both adjusted and unadjusted models, women married at age ≤14 were more likely to experience terminated or unintended pregnancy (AOR = 1.89, 95% CI: 1.31-2.75 and AOR = 2.20. 95% CI: 1.24-3.91, respectively), inadequate ANC (AOR = 1.71, 95% CI: 1.01-2.90), unmet need for family planning (AOR = 1.44, 95% CI: 1.05-1.98), fistula (AOR = 2.36, 95% CI: 1.22-4.57); While, for those married at age 15-17 years, only terminated or unintended pregnancy remained significant.

**Conclusion:** The younger age of marriage was associated with a higher prevalence of adverse reproductive outcomes among Afghan women. Moreover, poverty and illiteracy proved to be important predictors of child marriage in Afghan women. Strict international law enforcement and advocacy are a need in the current situation of Afghanistan to increase young women’s education, promote their civil rights, and improve their autonomy and role in decision-making concerning their health.

## Introduction

Child marriage is defined as marriage before the age of 18 years and is increasingly recognized as a violation of human rights and a serious public health issue (1). Although one-fifth of women worldwide were subjected to child marriage in 2020, the geographic distribution of child marriage could vary significantly across the regions and countries and it is mostly concentrated in poor- and middle-income countries (2). For instance, approximately a third of women in Sub-Saharan Africa were subjected to child marriage in 2017 (3); while this figure for South Asia was about 45% the same year with different rates across countries in this region (4). Child marriage could not only harm the gender equality efforts but also the achievement of millennium development goals, particularly goals 1 to 5: eradication of poverty, achievement of universal education, women empowerment, reducing child mortality, and improving maternal health (5).

It has been shown that it is associated with a number of adverse health and social outcomes such as unintended and high-risk pregnancy, maternal and infant mortality, human immunodeficiency virus/acquired immunodeficiency syndrome, cross-generational sex, obstetric fistula, high maternal mortality and morbidity, intimate partner violence, and depression (6-8). These adverse health outcomes have been attributed to several factors, including limited access to media and inadequate health education, less autonomy in decision making and inability to actively engage in health promotion programs, limited social engagement, and inability to adequately access health care (9, 10). Empowering young adolescent girls is essential to improve the overall reproductive and child health outcomes and accelerate social and economic development in low- and middle-income countries (11). However, it appeared that the content of reproductive health programs is often not serving this key vulnerable population who may be at a higher risk for poor reproductive health outcomes (1, 12).

Despite the current evidence of possible adverse reproductive outcomes among girls who marry at early ages, there is still a gap in knowledge of true association and determinants of child marriage and reproductive outcomes in some of the most afflicted countries such as Afghanistan. The long-standing ignorance of young women’s reproductive needs in patriarchal societies such as Afghanistan not only harmed the young women’s health but also the health of offspring who may suffer from perinatal complications (13). In addition, it limited the efforts of national and international parties in improving adolescent reproductive health through well-tailored programs that specifically target this vulnerable subgroup in Afghan society. To our knowledge, this is the first study in Afghanistan that explored the prevalence and determinants of child marriage and its association with adverse reproductive outcomes among a nationally representative sample of Afghan women. The results are to inform the policies and interventions of the most recent situation concerning child marriage and adverse reproductive outcomes in Afghanistan.

## Methods

### Study design

This was a cross-sectional study using the data from the most recent Demographic and Health Survey (DHS) in Afghanistan conducted in 2015. It is a nationally representative survey implemented by the Central Statistics Organization (CSO) in collaboration with the Afghanistan Ministry of Public Health (MoPH) and funded by the United States Agency for International Development (USAID).

### Study population and sampling

Afghanistan DHS 2015 (ADHS 205) is nationally representative that collected the data for women aged 15-49 years. The ADHS 2015 employed a stratified two-stage sample design to estimate the key indicators at the national level, in urban and rural areas, and for each of the 34 provinces of Afghanistan. At the first stage, 950 clusters consisting of EAs (enumeration areas) were selected; 260 in urban areas and 690 in rural areas. At the second stage, 25,650 households were selected through an equal probability systematic selection process. Weighting components were calculated and applied to obtain a representative estimate at the national level. All the women aged 15-49 years who were either permanent residents of the selected households or visitors who stayed in the households the night before the survey were recruited after informed consent. Our analyses have been restricted to women 20-24 years of age, the United Nations (UN) recommended age group for child marriage assessment (14). Selecting this age group not only enhances the comparability of the results but also minimizes the possibility of errors due to recall which may occur as the participants move to the older age groups (15).

### Scales and measurement

A standard DHS questionnaire including the questions related to the demographic and health issues was administered to all eligible women. The questionnaire collected data on women’s demographic characteristics, household information, family planning, fertility preferences, child health, sexually transmitted diseases, marriage and sexual behavior, and domestic violence (16). However, for the current study, our primary interest was questions related to child marriage and reproductive outcomes.

### Explanatory variables

The main explanatory variable was the age at first marriage which was translated into child marriage if the respondent’s age was less than 18 years at the time of marriage. The respondents were further classified into three groups based on the age at the first marriage: ≥18 years (marriage as an adult), 15-17 years (marriage in middle adolescence), and ≤14 years of age (marriage in early adolescence or childhood) to capture the potential vulnerability that may exist for the marriage in very early ages.

### Outcomes of interest

- ***Early fertility:*** It was a binary variable indicating whether the woman had a birth within the first 12 months of marriage.
- ***Unmet need for family planning:*** It was a binary variable that includes the unmet need for limiting (i.e. women whose most recent pregnancy was not wanted at all, fecund women who did not use contraception despite their desire to have no more children, women who were postpartum amenorrhoeic for 2 years following an unwanted birth and were not using contraception) and spacing (i.e. women whose most recent pregnancy was not wanted initially but wanted later, fecund women not using contraception who were undecided when/if they wanted a to have a child or who wanted a child 2+ years later, and women who were postpartum amenorrhoeic for 2 years following a mistimed birth and were not using contraception) (17).
- ***History of rapid repeat childbirth:*** It was defined as having had at least one birth within 24 months of previous childbirth.
- ***History of pregnancy termination:*** It was a binary variable indicating whether a woman has ever had a pregnancy ended by miscarriage, abortion, or stillbirth.
- ***Unintended pregnancy:*** It was a binary indicator of whether the respondent has had at least one child in the past 5 years that was wanted later or not wanted at all.
- ***Inadequate antenatal care (ANC):*** It indicates the number of antenatal visits during the last pregnancy. Based on the World Health Organization guidelines, less than 4 ANC visits were considered inadequate.
- ***Delivery by skilled personnel:*** It indicates whether the woman had received assistance from a skilled health care worker in her last birth.
- ***Institutional delivery:*** It was a binary variable indicating whether the woman delivered her last child in a health care facility (public or private).
- ***Fistula***: It was a binary variable defined as the involuntary loss of urine and/or feces through the vagina after labour.

### Covariates

Based on the literature review, the most relevant sociodemographic factors such as age (continuous in years), education (literate, illiterate), employment (yes/no) area of residence (rural, urban), wealth quintile (poor, middle, rich), husband age difference (≤5, 6-9, ≥10), husband education (literate, illiterate), polygamy, listening to radio, watching television, and ethnicity were also collected and accounted for in the multivariate analysis.

### Statistical Analysis

All the analyses were restricted to the age group 20-24 years. Descriptive statistics were employed to describe the respondent’s characteristics in this age group. Binary logistic regression estimated the odds of child marriage across the different sociodemographic factors. Multivariate Logistic regression models were constructed to examine the association between various outcome variables and age at marriage before and after adjustment for sociodemographic factors. Results are presented as odds ratio (OR) and adjusted odds ratio (AOR) with 95% confidence intervals (95% CI). To avoid the bias in estimates for the history of rapid repeat births, we restricted the sample to women who had a minimum of two children. All the analyses were performed using STATA version 14 and sampling weight and design were defined and accounted for using the STATA command “svy” in all analyses. All tests of hypothesis were 2-tailed, with a type 1 error rate fixed at 5%.

### Ethical consideration

The data used in this study could be accessed upon a reasonable request from the DHS website and are publicly available. The ethical necessities for data collection were sought and assured by the institutions that commissioned, funded, or managed the surveys. All DHS are reviewed and approved by ICF International and Institutional Review Board of Ministry of Health in the target country to ensure that the protocols comply with the U.S. Department of Health and Human Services regulations for the protection of human subjects. Therefore, this study did not require further ethical approval.

## Results

Table 1 presents the distribution of sociodemographic factors of the respondents, A total of 6083 young women aged 20-24 were included in this study to estimate the child marriage prevalence and explore its association with adverse reproductive outcomes in Afghan women living in Afghanistan. The majority of the respondents were illiterate (72.56%), unemployed (87.03%), and were living in rural areas (76.73%). Approximately 70% of the women were married to a man who was no more than 5 years older than them. Half of the husbands were literate and almost all of them were employed. Almost an equal proportion of women□a fifth□were in each wealth index group. Less than half of the respondents reported watching television (48.23%) or listening to the radio (42.31%) at least once in the last week. The highest proportion of respondents were from Pashtun (42.75%) and Tajik (32.70%) ethnic groups; while, the lowest proportions belonged to Baloch (0.52%), Nuristani (0.82%), and Pashai (0.96%).

**Table 1.** The characteristics of Afghan women aged 20-24 years, ADHS 2015 (N=6083)

| Variables | n (weighted%) |
| --- | --- |
| Age at marriage |  |
| $\geq 18$ (adult) | 1850 (47.97) |
| 15-17 (middle) | 1559 (38.92) |

### Child marriage prevalence and its sociodemographic determinants

As table 2 illustrates, almost half of the respondents married at ages less than 18 years, including 38.92% married as a middle adolescent (15-17 years of age) and 13.11% married as a young adolescent or a child (≤14 years of age). The odds of child marriage were 20% lower in literate women compared to illiterate ones. Women with ≥10 spousal age gap were more likely to endure child marriage compared to the reference group (spousal age gap ≤5). The women from the poorest households were more likely to marry at ages less than 18 years (OR = 1.43, 95% CI:1.09-1.89). The likelihood of early marriage (age <18) was significantly higher in Tajik (OR = 1.39, 95% CI:1.07-1.79) and Baloch (OR = 2.92, 95% CI:1.43-5.95) ethnic groups compared to reference group (Pashtun).

**Table 2.** The sociodemographic determinants of child marriage among Afghan aged 20-24 years, ADHS 2015

|  |  |
| --- | --- |
| adolescents) |  |
| $\leq 14$ (young adolescents/children) | 514 (13.11) |
| Age groups (years) |  |
| 20 | 1721 (26.74) |
| 21 | 736 (12.28) |
| 22 | 1278 (24.03) |
| 23 | 1144 (18.01) |
| 24 | 1104 (18.94) |
| Education |  |
| Illiterate | 4615 (72.56) |
| Literate | 1468 (27.44) |
| Employment |  |
| Employed | 769 (12.97) |
| Unemployed | 5314 (87.03) |
| Husband age gap (year) |  |
| $\leq 5$ | 4427 (70.17) |
| 6-9 | 985 (17.50) |
| $\geq 10$ | 671 (12.33) |
| Husband Education |  |
| Illiterate | 2926 (50.97) |
| Literate | 3987 (49.03) |
| Wealth index |  |
| Poorest | 1103 (19.29) |
| Poorer | 1388 (21.02) |
| Middle | 1307 (19.44) |
| Richer | 1323 (21.08) |
| Richest | 962 (19.17) |
| Place of Living |  |
| Urban | 1423 (23.27) |
| Rural | 4660 (76.73) |
| Watch television |  |
| Yes | 2685 (48.23) |
| No | 3384 (51.77) |
| Listen to radio |  |
| Yes | 2568 (42.31) |
| No | 3507 (57.69) |
| Ethnicity |  |
| Pashtun | 2715 (42.75) |
| Tajik | 1811 (32.70) |
| Hazara | 496 (8.64) |
| Uzbek | 384 (9.68) |
| Turkmen | 72 (1.91) |
| Nuristani | 323 (0.82) |
| Baloch | 68 (0.52) |
| Pashai | 113 (0.96) |
| Other | 91 (2.01) |

\* The total number for some variable does not add up to 6083 due to the missing data.

### The association between the age of marriage and adverse reproductive outcomes

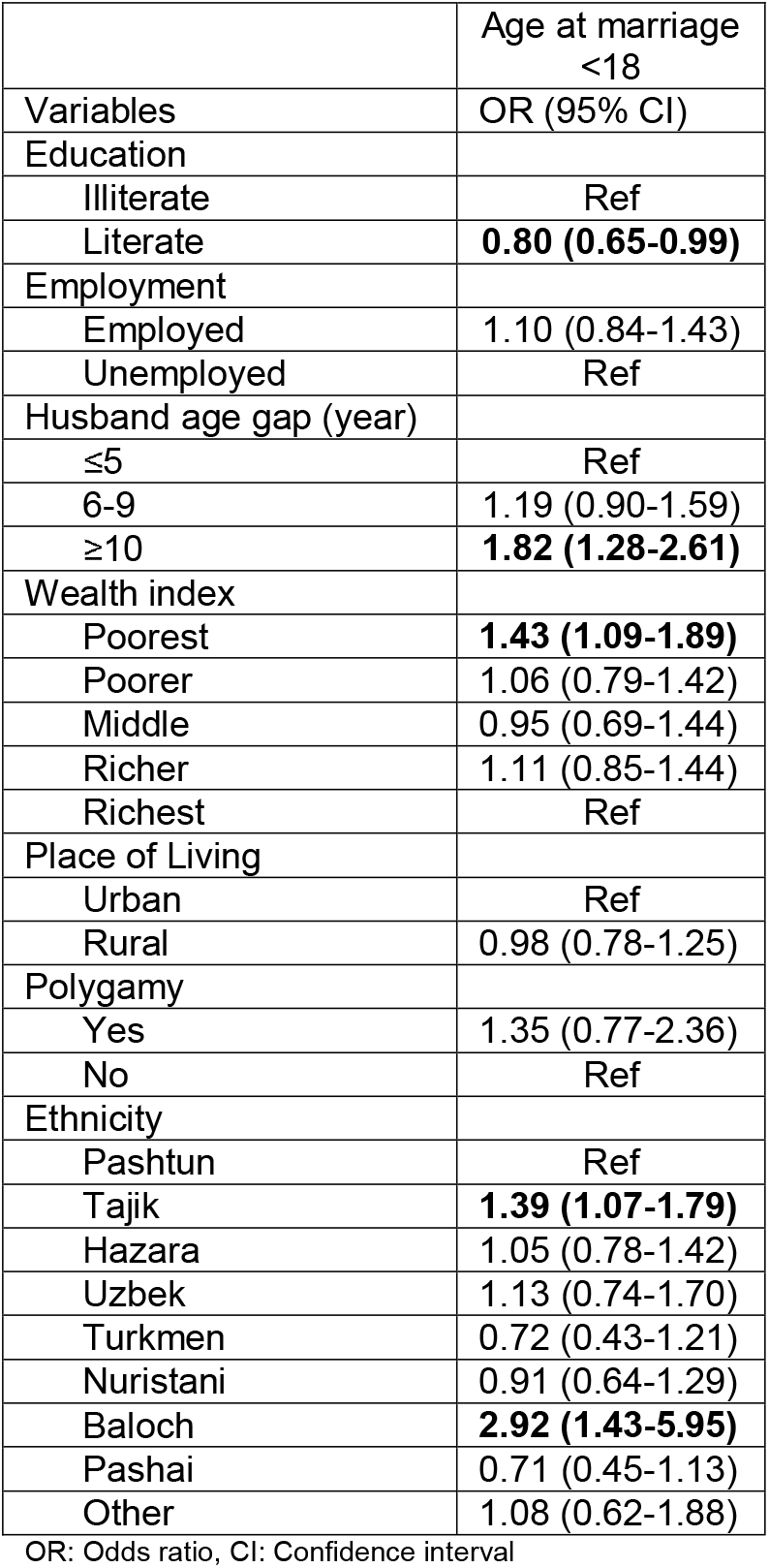

Table 3 represents the odds ratios (adjusted and unadjusted) and 95% confidence intervals for the association between child marriage and adverse reproductive outcomes. The age at marriage was classified into three categories: ≤14 (early adolescent/child), 15-17 (middle adolescent), and ≥18 (adult) to capture the variability of reproductive outcomes prevalence across the younger age groups. Nine reproductive outcomes were included in the analysis as below:

**Table 3.** The association between the age at marriage and adverse reproductive outcomes among Afghan women aged 20-24 years, ADHS 2015

|  |  |  |  |  |
| --- | --- | --- | --- | --- |
|  | Age at marriage<br>≤14<sup>a</sup> |  | Age at marriage<br>15-17<sup>a</sup> |  |
| Variables | OR (95% CI) | AOR<sup>b</sup> (95% CI) | OR (95% CI) | AOR<sup>b</sup> (95% CI) |
| Early fertility<sup>c</sup> | 0.90 (0.63-1.28) | 0.86 (0.60-1.24) | 0.94 (0.76-1.15) | 0.93 (0.75-1.16) |
| Unmet needs for family planning | **1.37 (0.99-1.90)** | **1.44 (1.05-1.98)** | 1.19 (0.93-1.54) | 1.23 (0.96-1.57) |
| History of rapid repeat childbirth<sup>d</sup> | **0.34 (0.24-0.48)** | **0.27 (0.18-0.39)** | **0.56 (0.42-0.74)** | **0.45 (0.33-0.61)** |
| History of pregnancy termination | **1.81 (1.26-2.59)** | **1.89 (1.31-2.75)** | 1.45 (0.98-2.13) | **1.57 (1.09-2.66)** |
| Unintended pregnancy | **1.94 (1.15-3.26)** | **2.20 (1.24-3.91)** | 1.32 (0.85-2.05) | **1.61 (1.03-2.56)** |
| Inadequate ANC (<4 ANC visits) | **1.92 (1.23-3.00)** | **1.71 (1.01-2.90)** | **1.55 (1.07-2.24)** | 1.42 (0.91-2.23) |
| Delivery by skilled personnel | **0.51 (0.38-0.69)** | **0.48 (0.34-0.69)** | **0.67 (0.53-0.86)** | **0.64 (0.48-0.85)** |
| Institutional delivery | **0.49 (0.36-0.66)** | **0.45 (0.32-0.66)** | **0.68 (0.54-0.86)** | **0.65 (0.49-0.85)** |
| Fistula | **2.24 (1.10-4.57)** | **2.36 (1.22-4.57)** | 1.39 (0.70-2.78) | 1.38 (0.69-2.75) |

OR: Odds ratio, CI: Confidence interval.
<sup>a</sup> The reference group: Age at marriage ≥18.
<sup>b</sup> AOR: Adjusted odds ratio (controlled for current age, education, wealth index, ethnicity, husband education, husband age gap, watching tv, listening radio).
<sup>c</sup> Within one year after marriage.
<sup>d</sup> Defined as birth interval < 24 months in last pregnancy.
\* Due to the small sample size of some ethnic groups, this variable was not included in multivariate analysis.

1. Early fertility: It appeared that there is no association between age at marriage and early fertility even after adjustment for sociodemographic factors.
2. Unmet need for family planning: Women married at ages ≤14 were more likely to suffer from the unmet needs for family planning not only before (OR = 1.37, 95% CI: 0.99-1.90) and but also after (AOR = 1.44, 95% CI: 1.05-1.98) adjustment for sociodemographic factors. No significant association was observed for those married at ages 15-17 years compared to the reference group (married at ≥18)
3. History of rapid repeat childbirth: It was defined as birth interval < 24 months in the last pregnancy and only women with at least two births were included. It appeared that the likelihood of having rapid repeat pregnancy is significantly lower among women married at both ≤14 and 15-17 years of age compared to those married at age >18 years, even after adjustment for sociodemographic factors (AOR = 0.27, 95% CI:0.18-0.39 and AOR = 0.45, 95% CI:0.33-0.61, respectively).
4. History of pregnancy termination (stillbirth, abortion, miscarriage): There was a significant positive association between child marriage and history of pregnancy termination after adjustment for sociodemographic factors. In the adjusted models, the odds of experiencing terminated pregnancy (stillbirth, abortion, or miscarriage) were almost 90% and 60% higher among those married at ≤14 and 15-17 years of age compared to those married at older ages.
5. Unintended pregnancy: The odds of experiencing unintended pregnancy among girls married at age ≤14 was almost twice the odds of women married at ≥18 in both unadjusted and adjusted models (OR = 1.94, 95% CI: 1.15-3.26 & AOR = 2.20, 95% CI:1.24-3.01). Likewise, in the adjusted model, women married at age 15-17 years were more likely to have an unintended pregnancy (AOR = 1.61, 95% CI: 1.03-2.56).
6. Inadequate ANC (<4 ANC visits): Women married at ages less than 18 were more likely to endure an inadequate ANC during their last pregnancy (OR = 1.92, 95% CI: 1.23-3.00 for women married at ≤14 and OR = 1.55, 95% CI: 1.07-2.24 for those married at 15-18); however, this association was weakened for those married at age 15-17 years after adjustment for sociodemographic factors (AOR = 1.42, 95% CI: 0.91-2.23).
7. Delivery by skilled personnel: it appeared that likelihood of having a delivery by a skilled health worker at last birth was significantly lower among women married at age <18. As Table 3 indicated, in both adjusted and unadjusted models, the odds of delivery by a skilled worker were almost 50% and 35% lower among women married at age ≤14 and 15-17 compared to those married at age >18 years, respectively.
8. Institutional delivery (public, private): Similar to the delivery by skilled personnel, the odds of having institutional delivery, in both adjusted and unadjusted models, was approximately 50% and 35% lower among women married at age ≤14 and 15-17 compared to those married at age >18 years, respectively.
9. Fistula: Women married at age ≤14 had significantly higher odds of experiencing involuntary loss of urine and/or feces through the vagina after labour compared to those married at age >18 even after adjustment for sociodemographic factors (OR: 2.24, 95% CI: 1.10-4.57 & AOR = 2.36, 95% CI: 1.22-4.57). However, for those married at age 15-17 years, no significant relation was observed

## Discussion

Although several studies examined the impact of child marriage on adverse reproductive outcomes in other countries and regions such as South Asian countries (4, 18, 19) or Sub-Saharan African countries (20, 21), to the best of our knowledge, this is the first study that explored such relationship among Afghan women living in Afghanistan. The results showed that an estimated 52% of Afghan women aged 20-24 married at ages less than 18 years. Furthermore, there was a significant association between marriage at early ages and adverse reproductive outcomes among young Afghan women living in Afghanistan. It appeared that women married at age ≤14 are more vulnerable and except for early fertility, they were more likely to experience any other type of adverse reproductive outcomes including the history of an unintended or terminated pregnancy, unmet need for family planning, inadequate ANC, delivery by an unskilled worker in a non-institutional place, and fistula; whereas, marrying at age 15-17 years was only associated with a history of unintended, or terminated pregnancy and delivery by an unskilled worker in a not-institutional place such as home after adjustment for sociodemographic factors. However, they may also have inadequate ANC (<4) during their pregnancy, even though, this association disappeared after accounting for the effect of sociodemographic factors. We also found that there is a higher prevalence of child marriage among poor, illiterate women who belong to the Tajik and Baloch ethnic groups.

Although more than half the women in this study endured child marriage, it appeared that the prevalence of child marriage significantly decreased over the period 2010 to 2015 (76% in DHS 2010 vs 52% in DHS 2015 in women aged 20-24 years). This may be due to the national and international campaigns and enormous efforts toward empowering the Afghan women that have been shaped during the last decade. However, women from lower socioeconomic classes with low literacy are still at a greater risk for child marriage. Similar to our study, several studies have reported poverty and illiteracy as risk factors for child marriage (22) and adverse fertility outcomes (23, 24). Poverty and illiteracy appear to limit the women’s autonomy in critical decisions and often exclude them from the decisions related to their marriage and reproductive needs (24). In fact, it has been suggested that retaining girls at the school for a longer period not only raise the age at the first marriage but also enhances the autonomy of the young girls through individual development (18). in Afghanistan, child marriage is often a measure of strengthening ties between rival families and tribes and is considered a peaceful solution to settle debts and disputes. Many poor families often sell their adolescent girls in exchange for large dowries from rich people and the husbands are usually much older (25). Similarly, in our study, a higher chance of child marriage was observed in those with ≥10 years spousal age gap. Almost all the countries with a high prevalence of child marriage are located in the developing world (3). Child marriage is a common phenomenon in rural areas where poverty and illiteracy are more dominant. Several studies indicated a higher prevalence of child marriage among women living in rural areas (26). However, our study found no significant difference in child marriage between rural and urban respondents, this could be due to the widespread poverty and illiteracy of Afghan women regardless of their place of living. More studies are suggested to scrutinize this issue among rural and urban habitants in Afghanistan.

Child marriage has been linked to several maternal and child adverse health outcomes such as low birth weight, sexually transmitted diseases (STDs), unintended pregnancy, abortion, stillbirth, maternal and child mortality (6, 8). There are also negative social impacts such as social isolation and domestic violence (27); however, the findings are inconsistent. The discrepancies in findings across studies have been attributed to the differences in the socio-cultural and economic status of the target society (28). In our study, the odds of all adverse reproductive outcomes, except early fertility and history of rapid repeat pregnancy, were higher in women married at age ≤14 compared to those married as an adult (age >18). We found no significant association between early fertility and age at the marriage, which was in line with previous studies from India, Nepal, and Pakistan (4); however, there was a negative association between the history of rapid repeat pregnancy and age at marriage across all child marriage groups. This has been attributed to the low fecundity at very young ages due to the physical and biological immaturity and probably low coital frequency (29). Several factors could influence the coital frequency at very young ages such as arranged marriage, delayed consummation, and living in an extended family which reduces the chance of intercourse and early fertility (29).

An increase in odds of unmet need for family planning was documented among those married at very young ages in the present study. This could partly justify by an increased desire to use modern contraceptives among women married at young ages to prevent further unintended pregnancies (20). The women married at very young ages often attain the desired number of children in their twenties and they may seek contraceptives to avoid further pregnancy. This could be a reason for higher needs for family planning at younger ages compared to those married at older ages who may still want more children (30, 31). The higher unmet need for family planning obviously leads to a higher prevalence of unintended pregnancy, which was observed among women married at age <18 in our study. Besides, unintended pregnancy often leads to negative pregnancy outcomes such as abortion, miscarriage, and stillbirth, especially at very young ages due to the anatomical and physiological immaturity of the body (32, 33). Moreover, consistent with our finding, it has been shown that young girls aged 10-15 years are more likely to suffer from obstetric fistula due to their small pelvises (34). Therefore, the reasons and determinants of unmet need for family planning services among women married at very young ages in Afghanistan should be a priority for future research.

We observed a negative association not only between delivery by a skilled worker at institutional facilities but also between adequate ANC with child marriage. Similar findings have been documented in other countries such as Pakistan, Nepal, and India (35-37). According to previous studies, in Islamic patriarchal societies such as Afghanistan women married at very young ages are more likely to suffer from spousal violence and often controlled by their husbands or in-laws (38, 39). This is mainly due to the low empowerment of women marrying as children and this could limit the autonomy in decision-making and access to reproductive and maternal health services (40-42). Due to the dependence and low status of child brides, they need to seek permission from the head of a family to visit health services (43), otherwise, they may encounter spousal violence and this limits their access to maternal healthcare services (44). This may also explain the high prevalence of other adverse reproductive outcomes in our study.

## Limitations

Although our study is the first study that measures the prevalence of child marriage and its association with adverse reproductive outcomes in a nationally representative sample of Afghan women aged 20-24 years, some cautions need to be considered in interpreting the results. The study used self-reported data that are subjected to recall and socially desirable biases. For example, they may be an error in reporting the age of respondent and age of marriage as there was no documented registration system for age. Furthermore, some adverse reproductive outcomes such as unintended or terminated pregnancy may be underreported. Also, pregnancy termination did not differentiate between miscarriage, abortion, or stillbirth and hence determining the relationship of each complication with child marriage was not possible. Moreover, we were not able to distinguish between induced and natural pregnancy termination. Due to the cross-sectional design of DHS surveys, the causal relationship between child marriage and adverse reproductive outcomes may not be assumed. However, since the reproductive outcomes occur following marriage, this assumption may not stand for this particular study. Last, the findings were limited to the age group 20-24 years in Afghanistan and cannot be generalized to other ages. However, the same age group of women has been recommended by the UN and used in other international publications concerning child marriage (19, 20, 45)

## Conclusion

This study not only contributes to the current knowledge related to the impact of child marriage on reproductive outcomes but also described the most recent situation of child marriage and its sociodemographic determinants among Afghan women. The findings underscore the critical need for policies and interventions to strive toward increasing the age of marriage among Afghan girls. Although in recent years, Afghanistan has made substantial efforts to reduce child marriage in face of incredible internal conflict, continued support from the international community and human rights advocates are essential as the recent worsening and political conflict could be a backslide to the progress that has been already made over the years. Strict international law enforcement and advocacy are a need in the current situation of Afghanistan to increase young women’s education, promote their civil rights, and improve their autonomy and role in decision-making concerning their health.

## Data Availability

The DHS questionnaire that collected the data in Afghanistan's demographic and health survey in 2015 could be reached at DHS official website (https://www.dhsprogram.com). The dataset that was used in this study could be available upon a reasonable request from the DHS website.

## Abbreviations

UN: United Nations
DHS: Demographic and Health Survey
CSO: Central Statistics Organization
ANC: Antenatal care
AOR: Adjusted odds ratio
MoPH: Ministry of Public Health
STD: Sexually transmitted disease

## Consent for publication

Not applicable.

## Authors’ contribution

OD wrote the research protocol and performed the data analysis and prepared the final draft. TN provided critical feedback and comments on the data analysis and results and help in preparing the final draft.

## Acknowledgment

We would like to thank the staff at Walailak University Institute of Research for their kind support and technical assistance in this research.

